# Arrhythmic Outcomes in Catecholaminergic Polymorphic Ventricular Tachycardia

**DOI:** 10.1101/2021.01.04.21249214

**Authors:** Sharen Lee, Jiandong Zhou, Kamalan Jeevaratnam, Ishan Lakhani, Wing Tak Wong, Ian Chi Kei Wong, Chloe Mak, Ngai Shing Mok, Tong Liu, Qingpeng Zhang, Gary Tse

## Abstract

**Introduction:** Catecholaminergic polymorphic ventricular tachycardia (CPVT) is a rare cardiac ion channelopathy. The aim of this study is to examine the genetic basis and identify predictive factors for arrhythmic outcomes in CPVT patients from Hong Kong.

**Methods:** This was a territory-wide retrospective cohort study of consecutive patients diagnosed with CPVT at public hospitals or clinics in Hong Kong. The primary outcome was spontaneous ventricular tachycardia/ventricular fibrillation (VT/VF).

**Results:** A total of 16 (mean presentation age=11±4 years old) patients were included. All patients presented at or before 19 years of age. Fifteen patients (93.8%) were initially symptomatic. Ten patients had both premature ventricular complexes (PVCs) and VT/VF, whereas one patient had PVCs without VT/VF. Genetic tests were performed in 14 patients (87.5%). Eight (57.1%) tested positive for the RyR2 gene. Seven variants have been described elsewhere (c.14848G>A, c.12475C>A, c.7420A>G, c.11836G>A, c.14159T>C, c.10046C>T and c.7202G>A). c.14861C>G is a novel RyR2 variant that has not been reported outside this cohort. All patients were treated with beta-blockers, three patients received amiodarone and two received verapamil. Sympathectomy (n=8), ablation (n=1) and implantable-cardioverter defibrillator implantation (n=3) were performed. Over a median follow-up of 127 (IQR: 97-143) months, six patients suffered from incident VT/VF. No significant predictors were identified on Cox regression. Nevertheless, a random survival forest model identified initial VT/VF/sudden cardiac death, palpitations, QTc, initially symptomatic and heart rate as important variables for estimating the probability of developing incident VT/VF.

**Conclusion:** All CPVT patients who are from Hong Kong presented at or before 19 years of age. Clinical and electrocardiographic findings can be used to predict arrhythmic outcomes. A nonparametric machine learning survival analysis achieved high accuracy for predicting the probability of incident VT/VF.

## 1. Introduction

Cardiac ion channelopathies predispose to the development of spontaneous ventricular tachycardia/ fibrillation (VT/VF) and sudden cardiac death (SCD) [1-6]. Of these, catecholaminergic ventricular tachycardia (CPVT) is less prevalent conditions compared to Brugada syndrome (BrS) in Asia [7, 8]. It is typically caused by mutations in either the ryanodine receptor 2 (RyR2) [9] or the calsequestrin 2 (CASQ2) [10, 11], but mutations in other genes such as calmodulin (CALM) have been implicated [12-14]. CPVT is usually precipitated by exercise or distress, which results in bidirectional VT, presenting in the first two decades of life [15]. Globally, population-based data on CPVT have mainly come from Westrn countries. The largest registry created by the Pediatric and Congenital Electrophysiology Society of the United States reported the characteristics of 237 patients [16, 17]. In another multi-national study including mainly patients from France, outcomes in 101 patients were reported [18], complementing smaller registry and case series studies by the same group [19, 20]. Another study reported specifically 21 CPVT patients caused by the CALM genes [12].

By contrast, data from Asia have been relatively sparse. A multi-centre Japanese registry of 78 patients found that 94% of the cases were sporadic with only 6% of the cases being familial [21]. In a national study from Japan, it was found that 30 gene mutation carriers were found for 3 genes in 50 probands [22]. Another Japanese reported on the findings of 29 patients [23]. Two studies from China has reported on six patients [11] and 12 patients [24].

Machine learning methodologies have been applied to improve risk prediction for cardiovascular diseases [25], including arrhythmia research [26-29]. Of these, random survival forest (RSF) builds hundreds of trees and produces prediction of outcomes by a voting method for analysing right censored survival data [30]. Its advantages include a lack of assumption regarding the individual hazard function [31] and the ability to capture latent and complex interactions among the variables [31]. In this study, we investigated the genetic basis and risk factors for predicting arrhythmic outcomes using RSF in CPVT patients who presented to the public hospitals or clinics of Hong Kong, China.

## 2. Methods

### 2.1. Study Population

This study was approved by The Joint Chinese University of Hong Kong – New Territories East Cluster Clinical Research Ethics Committee. The cohort included consecutive patients with an initial diagnosis of CPVT between January 1^st^, 1997 to September 1^st^, 2019 in public hospitals or clinics. Our team has previously used this system for conduct disease-based epidemiological or clinical studies in patients with both prevalent and rare conditions [32, 33]. Centralized electronic health records were reviewed for patient identification and data extraction. The diagnosis was made initially by the case physicians. They were confirmed by G.T. and S.L. through the review of case notes, documented ECGs, diagnostic test results, and genetic reports. Diagnosis of CPVT was established based on the exercise treadmill test, adrenaline challenge test, or genetic testing by the participating institution at the time of entry.

### 2.2. Clinical and Electrocardiographic Data Collection

The baseline clinical data extracted from the electronic health records include: 1) sex; 2) age of first characteristic ECG presentation and last follow-up; 3) follow-up duration; 4) family history of SCD and the specific ion channelopathy; 5) syncope manifestation and its frequency; 6) presentation of sustained VT/VF and its frequency; 7) performance of electrophysiological study (EPS), 24-hours Holter study, ion channelopathy-specific genetic testing, and the respective results; 8) performance of echocardiogram; 9) presence of other arrhythmias; 9) implantation of implantable cardioverter-defibrillator (ICD); 10) occurrence, cause and age of death; 11) period between the initial presentation of characteristic ECG and the first post-diagnosis VT/VF episode; 12) initial disease manifestation (asymptomatic, syncope, VT/VF). In the present study, symptoms refer to syncope or VT/VF, thus asymptomatic indicates freedom from either presentation. Other arrhythmias include sick sinus syndrome, bradycardia, atrioventricular block, atrial tachyarrhythmias, and supraventricular tachyarrhythmias. Positive EPS is defined as the induction of spontaneous VT/VF that either sustained a minimum of 30 seconds or produced hemodynamic collapse.

The following automated measurements were extracted from baseline ECGs: 1) heart rate; 2) P wave duration (PWD) and PR interval; 3) QRS duration; 4) QT and QTc interval; 5) P, QRS and T wave axis; 6) amplitude of R and S wave from leads V5 and V1 respectively; 7) presence of 1^st^ degree atrioventricular block, defined as PR-interval greater than 200ms; 8) presence of interventricular delay, defined as QRS-interval greater or equal to 110ms. Baseline ECG is the documented ECG taken at or the earliest after the initial characteristic ECG presentation. All ECG parameters, except for the amplitude of R- and S-wave from leads V5 and V1 respectively, were averaged across the 12 leads.

### 2.3. Statistical Analysis

Categorical variables were compared using Fisher’s exact test and reported as total number (percentage), whilst discrete and continuous variables were compared by Kruskal-Wallis one-way ANOVA and expressed as mean±standard deviation. The mean annual VT/VF incidence rate of each subgroup is calculated by first obtaining the patient-specific rate through dividing the total number of sustained VT/VF events by the follow-up period, then average the rates within the subgroup. Statistical significance was defined as P-value < 0.05. The difference in the duration of post-diagnosis VT/VF-free survival between the pediatric and adult subgroup is compared quantitatively by Kaplan-Meier survival curve and compared using the log-rank test. All statistical analysis was performed using R Studio (Version: 1.3.1073).

### 2.4. Development of a machine learning survival analysis model

Survival analysis models were used to predict the risk of future time-to-events. Commonly, the default choice is Cox regression because of its convenience. Random Survival Forest (RSF) is a class of survival analysis models that use data on the life history of patients (the outcome or response) and their meaningful characteristics (the predictors or variables) [31]. RSF extends traditional random forests algorithm for a target which is not a class, or a number, but a survival curve. RSF is non-parametric and does not assume proportional risks as in the Cox model. This allows direct learning of the survival patterns between predictors and outcome. RSF bypasses the traditional necessity to impose parametric or semi-parametric assumptions on the underlying distributions of censored data and therefore provides an alternative approach to automatically deal with high-level interactions and higher-order nonlinear terms in variables and achieve much higher accurate survival predictions. The time-to-event prediction task through the RSF model aims to characterize the covariate effects on the time of a future VT/VF event, while capitalizing on meaningful information from censored data when performing learning.

A variable importance ranking approach was adopted based on standard bootstrap theory to investigate the strength of the associated significant univariate variables to predict VT/VF. Out-of-bag (OOB) method was adopted whenever a bootstrap sample is down with replacement from the training dataset. The importance value for the variable of interest was calculated as the prediction error (squared loss) for the original ensemble event-specific cumulative probability function subtracted from the prediction error for the new ensemble obtained using randomizing assignments of the variable [30]. Variable that were important predictors of VT/VF has a larger importance value, indicating higher predictive strength, whereas non-predictive variables have zero or negative values. Application of RSF model for predicting VT/VF were conducted using the *ggRandomForests* R package. Survival estimates were calculated using the Brier score (0=perfect, 1=poor, and 0.25=guessing) based on the inverse probability of censoring weight (IPCW) method [34]. The cohort was stratified into four groups of 0-25, 25-50, 50-75 and 75-100 percentile values of VT/VF.

## 3. Results

### Baseline characteristics, treatment and predictors identified using machine learning survival analysis

This study included 16 consecutive patients (mean presentation age= 11±4 years old; female= 50%; mean follow-up period= 116±36 months), all of whom presented below the age of 25 years old (**Tables 1 to 3**). Twelve patients fulfilled at least two criteria and four patients fulfilled one criterion of the 2013 HRS/EHRA/APHRS expert consensus statement (**Supplementary Table 1**). All patients were of Han Chinese origin. Of the whole cohort, 15 (93.8%) were initially symptomatic at presentation (i.e. patients with an initial presentation of syncope or VT/VF). Ten patients had both PVCs and VT/VF, whereas one patient had PVCs without VT/VF. The median delay between initial presentation and diagnosis was 4.5 (Q1-Q3: 0.8-8.5) months. Genetic tests were performed in 14 patients (87.5%), of which 8 patients (57.1%) tested positive for gene mutations. All mutations involved the RyR2 gene (**Supplementary Table 2**). The c.14861C>G mutation is novel and have not been described beyond our locality.

**Table 1.** Baseline clinical and demographic characteristics. Categorical and continuous variables were compared between groups using Fisher’s exact test or t-test, respectively. Bolded text indicate P<0.05.

| Variable | All CPVT patients (n=16) | CPVT patients with incident VT/VF on follow-up (n=6) | CPVT patients without incident VT/VF on follow-up (n=10) | P-value |
| --- | --- | --- | --- | --- |
| Female | 8 (50.0) | 4 (66.7) | 4 (40.0) | 0.302 |
| Presentation Age (years) | 10.8±4.4 | 10.7±4.1 | 10.8±4.9 | 0.478 |
| Diagnosis Age (years) | 11.4±4.4 | 11.3±3.8 | 11.4±5.0 | 0.489 |
| Current Age (years) | 20.5±6.5 | 21.8±3.4 | 19.7±7.9 | 0.729 |
| Presentation to Diagnosis (months) | 7.7±10.4 | 7.5±6.8 | 7.8±12.4 | 0.479 |
| Family History of CPVT/SCD | 3 (18.8) | 3 (50.0) | 0 (0) | 0.137 |
| Initially symptomatic | 15 (93.8) | 6 (100.0) | 9 (90.0) | 0.424 |
| Initial syncope | 14 (87.5) | 5 (83.3) | 9 (90.0) | 0.696 |
| Initial VT/VF/SCD | 5 (31.3) | 1 (16.7) | 4 (40.0) | 0.330 |
| Initial palpitations | 4 (25.0) | 3 (50.0) | 1 (10.0) | 0.074 |
| Initial chest pain | 1 (6.3) | 1 (16.7) | 0 (0) | 0.182 |
| Initial seizure | 6 (37.5) | 3 (50.0) | 3 (30.0) | 0.424 |
| PVC | 11 (68.8) | 5 (83.3) | 6 (60.0) | 0.330 |
| VT/VF | 10 (62.5) | 6 (100.0) | 4 (40.0) | **0.016** |
| VT/VF post-presentation | 6 (37.5) | 6 (100.0) | 0 (0) | - |
| Follow-Up Duration (months) | 116.3±35.9 | 131.7±17.5 | 107.0±41.5 | 0.904 |

**Table 2.** Results of different investigations. Categorical and continuous variables were compared between groups using Fisher’s exact test or t-test, respectively. Bolded text indicate P<0.05.

| Variable | All CPVT patients (n=16) | CPVT patients with incident VT/VF on follow-up (n=6) | CPVT patients without incident VT/VF on follow-up (n=10) | P-value |
| --- | --- | --- | --- | --- |
| **Cardiac imaging data** |  |  |  |  |
| Echocardiogram | 15 (93.8) | 5 (83.3) | 10 (100) | 0.182 |
| Abnormal echocardiogram | 1 (6.7) | 0 (0) | 1 (10.0) | 0.464 |
| Cardiac MRI performed | 5 (31.3) | 2 (33.3) | 3 (30.0) | 0.889 |
| Abnormal cardiac MRI | 0 (0) | 0 (0) | 0 (0) | 1.00 |
| **Data from other tests and clinical studies** |  |  |  |  |
| Genetic Test | 14 (87.5) | 5 (83.3) | 9 (90.0) | 0.696 |
| Positive Genetic Test | 8 (57.1) | 2 (40.0) | 6 (66.7) | 0.207 |
| Adrenaline Challenge | 7 (43.8) | 2 (33.3) | 5 (50.0) | 0.515 |
| Positive Adrenaline Challenge | 7 (100) | 2 (100.0) | 5 (100.0) | 1.00 |
| Exercise Tolerance Test | 15 (93.8) | 6 (100.0) | 9 (90.0) | 0.424 |
| Positive Exercise Tolerance Test | 13 (92.9) | 5 (83.3) | 8 (88.9) | 0.231 |
| EPS | 2 (12.5) | 1 (16.7) | 1 (10.0) | 0.696 |
| Positive EPS | 2 (100) | 1 (100.0) | 1 (100.0) | 1.00 |
| Holter Study | 14 (87.5) | 5 (83.3) | 9 (90.0) | 0.696 |
| Arrhythmia in Holter Study | 6 (46.2) | 4 (80.0) | 2 (22.2) | 0.053 |
| EEG | 8 (50) | 5 (83.3) | 3 (30.0) | **0.039** |
| Positive EEG | 0 (0) | 0 (0) | 0 (0) | 1.00 |

**Table 3.**
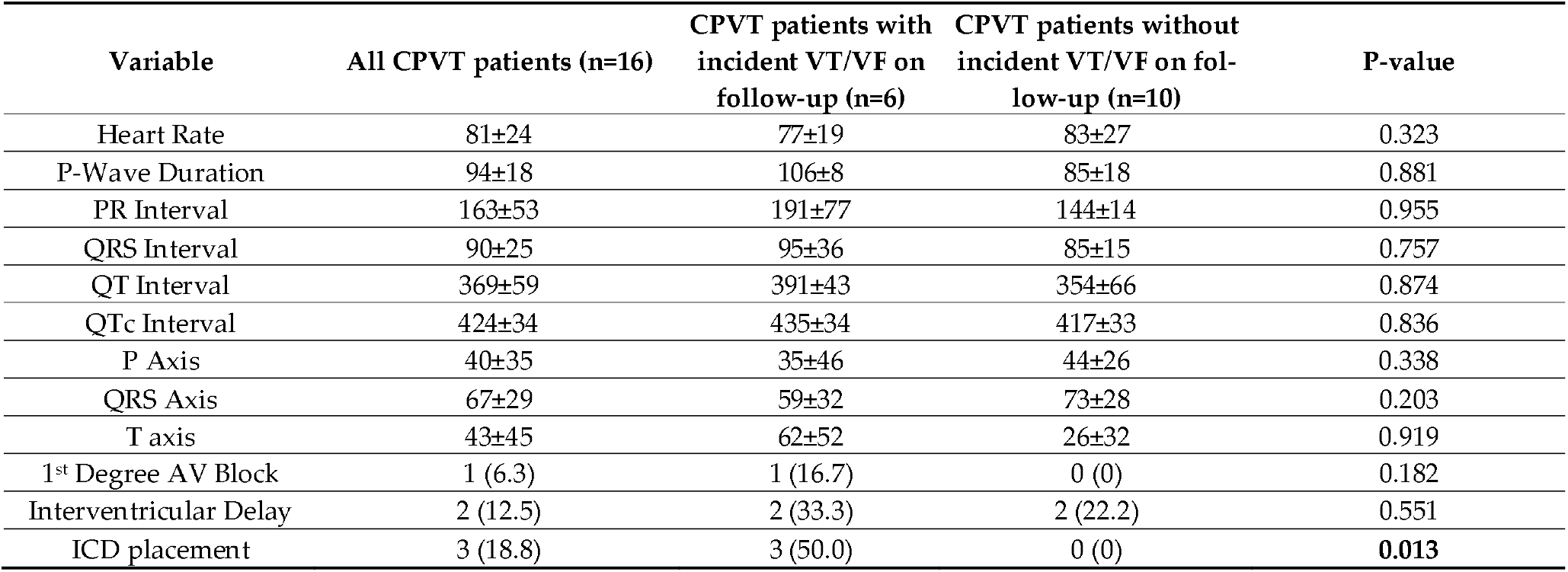
Baseline electrcardiographic data. Categorical and continuous variables were compared between groups using Fisher’s exact test or t-test, respectively. Bolded text indicate P<0.05.

| Variable | All CPVT patients (n=16) | CPVT patients with incident VT/VF on follow-up (n=6) | CPVT patients without incident VT/VF on follow-up (n=10) | P-value |
| --- | --- | --- | --- | --- |
| Heart Rate | 81±24 | 77±19 | 83±27 | 0.323 |
| P-Wave Duration | 94±18 | 106±8 | 85±18 | 0.881 |
| PR Interval | 163±53 | 191±77 | 144±14 | 0.955 |
| QRS Interval | 90±25 | 95±36 | 85±15 | 0.757 |
| QT Interval | 369±59 | 391±43 | 354±66 | 0.874 |
| QTc Interval | 424±34 | 435±34 | 417±33 | 0.836 |
| P Axis | 40±35 | 35±46 | 44±26 | 0.338 |
| QRS Axis | 67±29 | 59±32 | 73±28 | 0.203 |
| T axis | 43±45 | 62±52 | 26±32 | 0.919 |
| 1<sup>st</sup> Degree AV Block | 1 (6.3) | 1 (16.7) | 0 (0) | 0.182 |
| Interventricular Delay | 2 (12.5) | 2 (33.3) | 2 (22.2) | 0.551 |
| ICD placement | 3 (18.8) | 3 (50.0) | 0 (0) | **0.013** |

All patients were treated with beta-blockers (nadolol: n=12, metoprolol: n=5, atenolol: n=3, propranolol: n=3, sotalol: n=2), three patients received amiodarone and two received verapamil. Regarding invasive treatments, sympathectomy was performed in eight patients, ablation was performed in one patient and implantable-cardioverter defibrillators were inserted in three patients.

Over a median follow-up of 127 (IQR: 97-143) months, six patients suffered from incident VT/VF. The Kaplan-Meier post-diagnosis VT/VF-free survival curve is shown in **Figure 1a**. The application of RSF yielded the importance ranking of significant risk predictors for adverse outcome. The optimal tree number of RSF to predict VT/VF was set to 100 for RSF to predict VT/VF of CPVT through tree number iteration (**Figure 1b**). For CPVT, the most predictive five variables in predicting incident VT/VF were initial VT/VF or sudden cardiac death, an initial presentation with palpitations, QTc interval, initially symptomatic and heart rate (**Table 4**). The predicted OOB survivals and cumulative hazards with the RSF model to predict VT/VF in are shown in **Supplementary Figure 1**, which demonstrates efficient and accurate prediction strength of RSF model for incident VT/VF.

**Figure 1.**
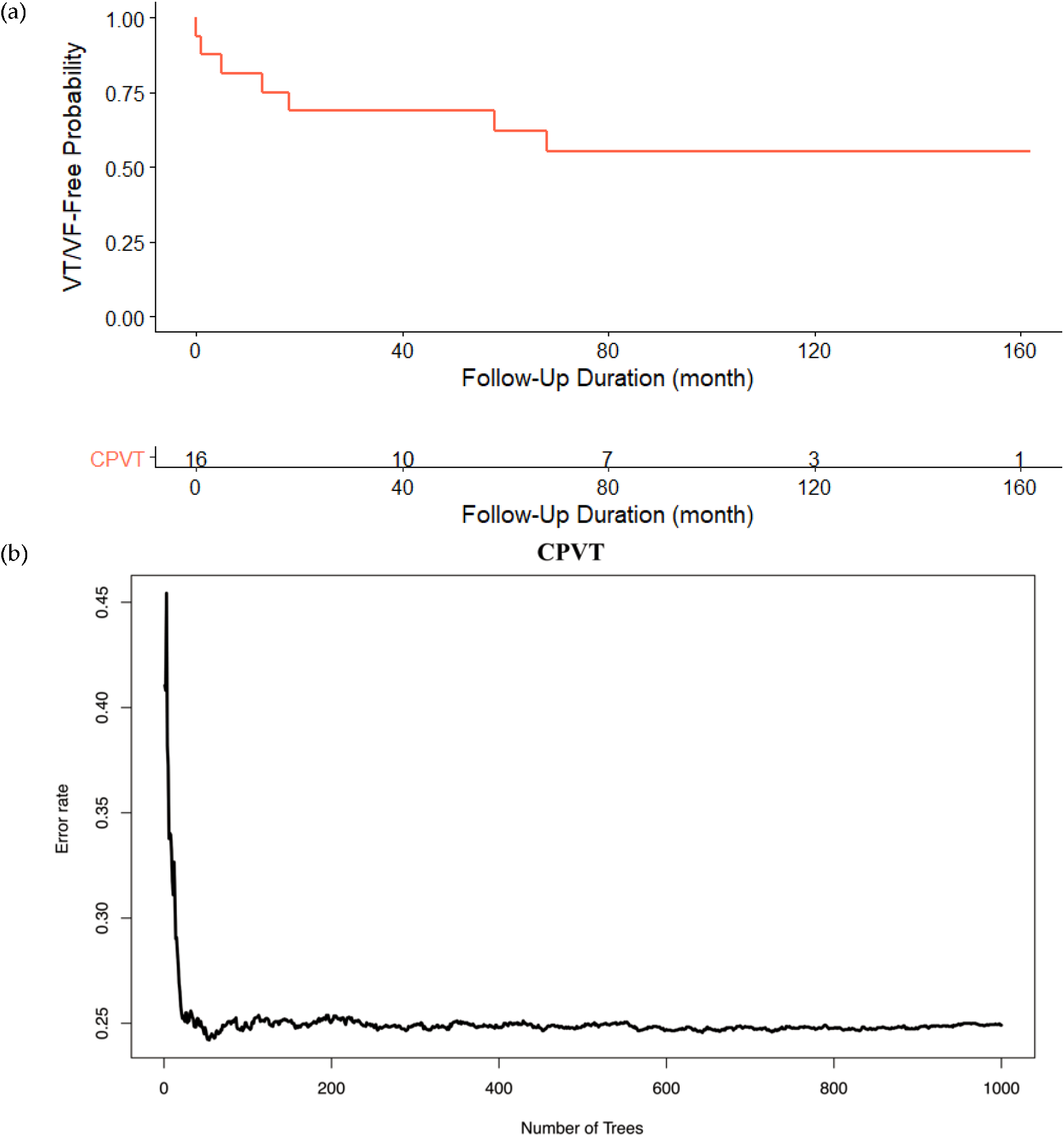
Kaplan-Meier survival curve for catecholaminergic polymorphic ventricular tachycardia (CPVT) patients (a). Optimal tree number selection for RSF model (*b*).

**Table 4.** Top five important variables for predicting incident VT/VF in CPVT generated by the RSF model.

| Predictor | Variable importance | Rank |
| --- | --- | --- |
| Initial VT/VF | 0.1033 | 1 |
| Initial palpitations | 0.0415 | 2 |
| QTc | 0.0307 | 3 |
| Initially Symptomatic | 0.0051 | 4 |
| Heart Rate | 0.0028 | 5 |

The survival functions estimated for each patient using the RSF model to predict VT/VF are shown in **Supplementary Figure 2a-d**. Specifically, the overall ensemble survival is indicated by the red line, whereas the Nelson-Aalen estimator is given by the green line *(a)*. The Brier score (0=perfect, 1=poor, and 0.25=guessing) stratified by ensemble VT/VF based on the inverse probability of censoring weight (IPCW) method shown in *(b)*. The cohort was stratified into four groups of 0-25, 25-50, 50-75 and 75-100 percentile VT/VF (the overall, non-stratified, Brier score is shown by the red line). A continuous rank probability score (CRPS) given by the integrated Brier score divided by time is shown in *(c)*, whereas a plot of VT/VF of each individual versus observed time is shown in *(d)*. Events are shown as blue points, whereas censored observations are shown as red points. The predicted VT/VF survivals by the RSF model are shown in **Figure 2**. The blue curves correspond to censored observations experiencing VT/VF events while the red curves correspond to observations experiencing VT/VF events predicted by using RSF model. The results demonstrated efficient performance of RSF model to generate well-fitted and accurate predictions of the adverse VT/VF events.

**Figure 2.**
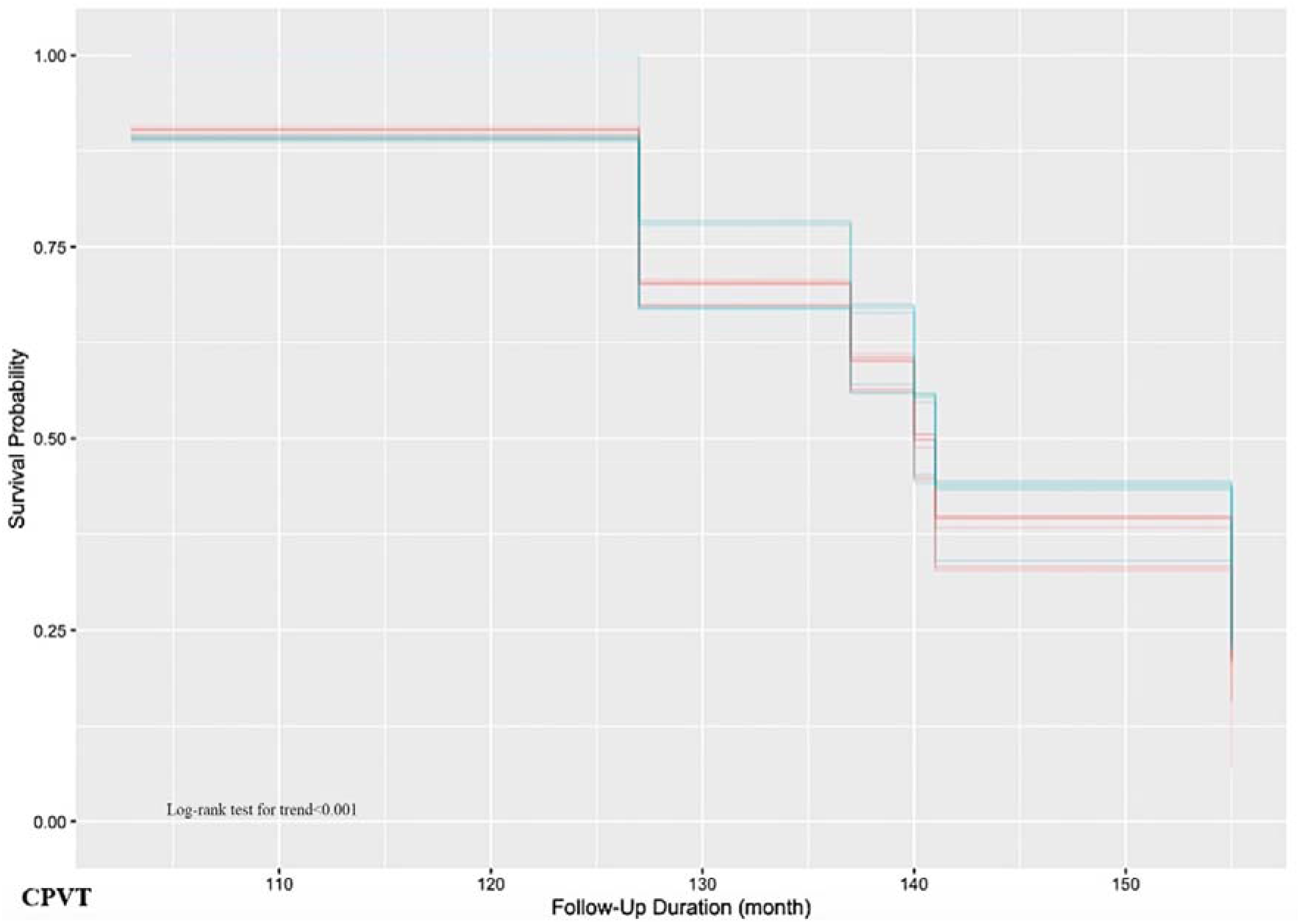
Predicted VT/VF survivals of CPVT patients generated by the RSF model. The blue curves correspond to censored observations experiencing VT/VF events while the red curves correspond to observations experiencing VT/VF events predicted by using RSF model.

## 4. Discussion

This is the first territory-wide cohort study of CPVT patients from Hong Kong, China. There are several major findings for the present study: 1) significant risk factors for spontaneous ventricular arrhythmias are initial VT/VF or sudden cardiac death, an initial presentation with palpitations, QTc interval, initially symptomatic and heart rate, 2) a nonparametric machine learning survival analysis is capable of automatic direct learning, with the ability to consider high-level interactions and higher-order nonlinear terms, thereby enabling the accurate prediction of incident VT/VF.

Sudden cardiac death is an important clinical problem globally, with congenital and acquired causes [35-38]. Of the congenital cardiac ion channelopathies, CPVT is characterized by exercise-induced bidirectional VT. International registry studies on European and North American patients have reported that there is a malignant arrhythmic phenotype associated with this disease with significant delays between initial presentation and subsequent diagnosis of around six months [17, 39]. By contrast, the epidemiology and characteristics of studies in Asia are limited. Recently, we have examined sudden arrhythmic death syndrome in young patients, but identified only two cases of CPVT locally [40]. In this multi-center study, we reported the findings of 16 patients, confirming a highly arrhythmic phenotype, with 10 patients with VT/VF at presentation or on follow-up. The application of RSF was able to identify the most important clinical and electrocardiographic predictors, which are in line with those reported by existing registries. Previously, RSF has been reported to enhance the model’s performance for predicting sudden arrhythmic deaths in patients with left ventricular ejection fraction <=35% [41], ischaemic heart disease [42] and ventricular tachyarrhythmias in congenital long QT syndrome [43], acquired long QT syndrome [44] and Brugada syndrome [45].

Several studies have examined the occurrence of adverse outcomes in CPVT cohorts, with particular emphasis on syncopal events and SCD [54-57]. There is existing evidence to suggest that subjects who are initially symptomatic, as similarly shown in our study, as well as those who are younger at diagnosis and are not administered β-blocker therapy have a significantly higher risk of cardiac events, including syncope, aborted cardiac arrest, and/or sudden cardiac death [55]. Likewise, findings indicate that an initial symptomatic presentation and an absence of β-blocker administration have also shown to be associated with mortality in CPVT patients [55]. Regarding electrocardiographic parameters, there is a relative paucity in literature assessing their use in risk prediction for VT/VF in the setting of CPVT. However, in the context of SCD as outcome, despite the fact that some reports studying its relationship with ECG variables have demonstrated significant differences in the QRS duration of recorded PVCs between patients who remained alive and those who suffered SCD during follow-up, most other ECG variables, such as those investigated in our study, namely heart rate and QTc interval, failed to demonstrate any notable variations with time [58].

Regarding the genetic basis, this study identified eight variants. Of these, c.14848G>A [46], c.12475C>A [47], c.7420A>G [48], c.11836G>A [49], c.14159T>C, c.10046C>T[50, 51] and c.7202G>A [52] have been reported elsewhere. By contrast, c.14861C>G is a novel RyR2 variant that gives rise to the A4954G amino acid change. This mutation affects the cytoplasmic domain of the RyR2, is expected to produce abnormalities in calcium handling, possible diastolic cacium leak and triggered arrhythmogenesis [53]. However, functional studies are needed to determine the precise mechanisms by which this structural change can lead to the generation of electrophysiological substrate. Previous animal studies have reported that the RyR2 mutations can be associated with not only disrupted calcium homeostasis but also reduced conduction velocity [59-61].

### Strengths and limitations

The major strengths of the present study include 1) predictors of post-diagnosis VT/VF free survival were derived for adult and pediatric/young patients; 2) holistic differences in clinical and ECG aspects of adult and pediatric/young patients were evaluated; 3) the study cohort was followed-up for a substantial length of time.

Several limitations should be noted for the present study. First, the retrospective nature of the study is inherently subjected to selection and information bias as well as missing data. However, consultations were performed at least annually for most patients, hence the patients were closely followed-up. Secondly, the predictive value of predictors is limited by the relatively small sample size as CPVT is the rarest out of the different ion cannelopathies in our city. Finally, given a lack of significance of the different input variables on Cox regression, it was not possible to compare the performance of RSF with Cox regression.

## 5. Conclusion

All CPVT patients from Hong Kong presented at or before 19 years of age. Several clinical and electrocardiographic findings were identified as important predictors of VT/VF, identified accurately through non-machine learning survival analysis. Specifically, the most important predictors determined include the presence of initial VT/VF, initial palpitations, initial symptomatic presentation as well as heart rate and QTc interval. Collectively, such parameters should be considered in clinical settings for the purposes of risk stratification and prognostic assessment in order to guide management and prevention of adverse arrhythmias in CPVT patients.

### Contributor statement

Sharen Lee and Gary Tse: study conception, data acquisition, database building, statistical analysis, manuscript drafting, manuscript revision Kamalan Jeevaratnam, Jiandong Zhou, Ishan Lakhani, Wing Tak Wong, Ian Chi Kei Wong, Chloe Mak, Ngai Shing Mok, Tong Liu, Qingpeng Zhang: data interpretation, statistical analysis, manuscript revision

### Institutional Review Board Statement

The study was conducted according to the guidelines of the Declaration of Helsinki, and approved by The Joint Chinese University of Hong Kong – New Territories East Cluster Clinical Research Ethics Committee (protocol number 2019.338, date of approval: 10^th^ September 2019; 2019.361, date of approval: 15^th^ August 2019).

### Informed Consent Statement

Patient consent was waived due to the retrospective and observational nature of this study.

## Supporting information

Anonymised dataset

## Data Availability

The anonymised data produced are available online at https://zenodo.org/record/5636292

## Data Availability Statement

Deidentified data are available upon request to the corresponding author.

