## Supplementary Appendix for "Arrhythmic Outcomes in Catecholaminergic Polymorphic Ventricular Tachycardia"

**Supplementary Table 1.** Details on the diagnostic criteria met for individual CPVT patients. The criteria proposed by the 2013 HRS/EHRA/APHRS expert consensus statement were used:

1. CPVT is diagnosed in the presence of a structurally normal heart, normal ECG, and unexplained exercise or catecholamine-induced bidirectional VT, polymorphic ventricular premature beats or VT in individuals <40 years of age.
2. CPVT is diagnosed in patients (index case or family member) who have a pathogenic mutation.
3. CPVT is diagnosed in family members of a CPVT index case with a normal heart who manifests exercise-induced PVCs or bidirectional/polymorphic VT.
4. CPVT can be diagnosed in the presence of a structurally normal heart and coronary arteries, normal ECG, and unexplained exercise or catecholamine-induced bidirectional VT, polymorphic ventricular premature beats or VT in individuals >40 years of age.

| Case number | Criteria 1 | Criteria 2 | Criteria 3 | Criteria 4 |
| --- | --- | --- | --- | --- |
| 1 | 1 | 1 | 0 | 0 |
| 2 | 1 | 1 | 0 | 0 |
| 3 | 1 | 1 | 0 | 0 |
| 4 | 1 | 1 | 0 | 0 |
| 5 | 0 | 1 | 1 (brother, mother, maternal granduncle) | 0 |
| 6 | 1 | 1 | 1 (brother, mother – not related to case 5) | 0 |
| 7 | 1 | 1 | 0 | 0 |
| 8 | 1 | 1 | 0 | 0 |
| 9 | 1 | 0 | 0 | 0 |
| 10 | 1 | 1 | 0 | 0 |
| 11 | 0 | 1 | 1 (father, sister) | 0 |
| 12 | 1 | 1 | 0 | 0 |
| 13 | 1 | 0 | 0 | 0 |
| 14 | 1 | 1 | 0 | 0 |
| 15 | 1 | 1 | 0 | 0 |
| 16 | 1 | 1 | 0 | 0 |

**Supplementary Table 2.** Genetic testing in individual CPVT patients. All mutations detected were in the RyR2 gene.

| Case number | Genetic test performed | Abnormal genetic test | Genetic results | Novel compared to overseas studies | Reference |
| --- | --- | --- | --- | --- | --- |
| 1 | 1 | 1 | c.14848G>A | No | (Priori et al., 2002) |
| 2 | 1 | 1 | c.12475C>A | No | (Kawata et al., 2016) |
| 3 | 1 | 0 | - | - | - |
| 4 | 1 | 1 | c.7420A>G | No | (Ozawa et al., 2018) |
| 5 | 1 | 0 | - |  |  |
| 6 | 1 | 1 | c.11836G>A | No | (Gallegos-Cortez et al., 2020) |
| 7 | 1 | 0 | - | - | - |
| 8 | 1 | 1 | c.14861C>G | Yes | - |
| 9 | 0 | - | - | - | - |
| 10 | 1 | 1 | c.14159T>C | No | RCV000182842 |
| 11 | 1 | 1 | c.10046C>T | No | (Christiansen et al., 2016;Seidelmann et al., 2017) |
| 12 | 1 | 0 | - | - | - |
| 13 | 0 | - | - | - | - |
| 14 | 1 | 0 | - | - | - |
| 15 | 1 | 0 | 0 | - | - |
| 16 | 1 | 1 | c.7202G>A | No | (Aizawa et al., 2005) |

**Supplementary Figures**

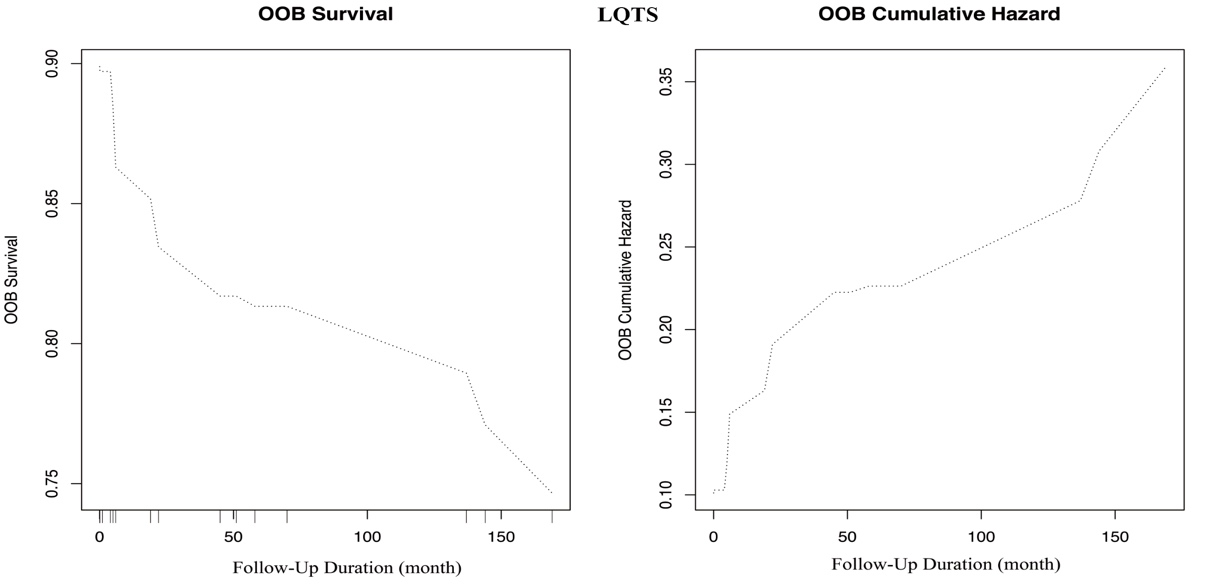

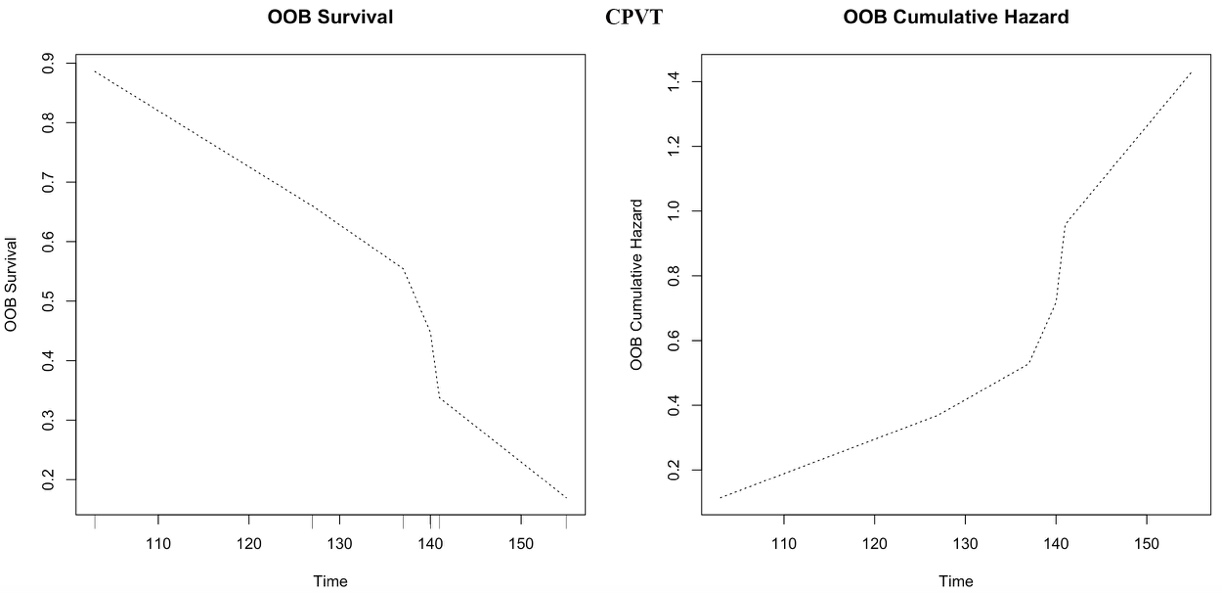

**Supplementary Figure 1. Predicted OOB survivals and cumulative hazards generated by the RSF model for predicting incident VT/VF in LQTS (*a, b*) and CPVT (*c, d*).**

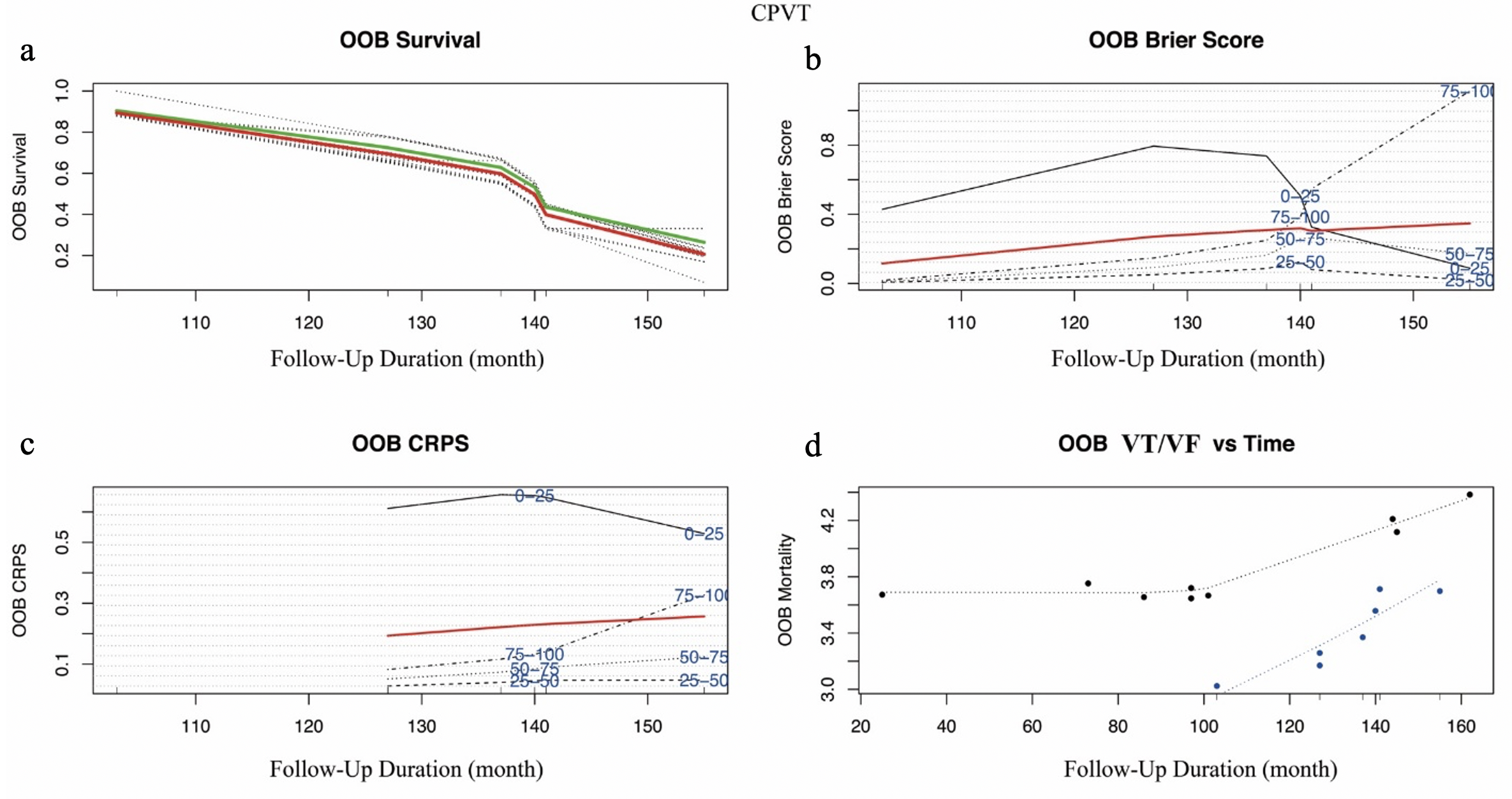

**Supplementary Figure 2. Survival estimates for the CPVT cohort generated from the random survival forest (RSF) model.** The overall ensemble survival is indicated by the red line; the Nelson-Aalen estimator is given by the green line (*a*). Brier score (0=perfect, 1=poor, and 0.25=guessing) stratified by ensemble mortality based on the inverse probability of censoring weight (IPCW) method (*b)* The cohort was stratified into four groups of 0-25, 25-50, 50-75 and 75-100 percentile mortality (the overall, non-stratified, Brier score is shown by the red line). Continuous rank probability score (CRPS) given by the integrated Brier score divided by time (*c*). The plot of incident VT/VF of each LQTS patient versus observed time (*d*). Events are shown as blue points, whereas censored observations are shown as red points.
